# The Effect of Intravenous Dexamethasone in Supraclavicular Brachial Plexus Block at Halibet National Referral Hospital, Eritrea 2024 - 2025: A Randomized Clinical Trial

**DOI:** 10.1101/2025.11.19.25340608

**Authors:** Michael Beraki Mengistu, Senay Amare Habtu, Abel Mussie Elias, Sabrina Hashim Idris, Kisanet Huruy Abraha

**Author notes:** **Contribution**: The Authors contributed equally to this work.

## Abstract

**Background:** Postoperative pain significantly impacts recovery and healthcare costs. Supraclavicular brachial plexus block (SCBPB) offers effective regional anesthesia but has limited duration. However, dexamethasone may enhance and prolong analgesia when used as an adjunct. Therefore, this study aimed to evaluate the effective of dexamethasone added to bupivacaine in prolonging postoperative analgesia.

**Method:** A randomized clinical trial was conducted at Halibet National Referral Hospital, Eritrea, involving 60 patients undergoing upper limb surgery. Group A received 0.5% bupivacaine (20 ml) with 2 ml normal saline; Group B received 0.5% bupivacaine (20 ml) with 2 ml (8 mg) intravenous dexamethasone. Pain scores, time to first rescue analgesia, and analgesic consumption were recorded. Data were analyzed using SPSS v26.

**Results:** The mean duration of analgesia was significantly longer in the dexamethasone group (17.36 ± 3.75 hours) compared to the control group (7.57 ± 1.04 hours, p < 0.001). The dexamethasone group had faster onset of sensory and motor block and lower analgesic consumption.

**Conclusions:** The addition of 8 mg intravenous dexamethasone to 0.5% bupivacaine in supraclavicular brachial plexus blocks significantly prolonged postoperative analgesia, accelerated the onset of sensory and motor block, and reduced analgesic consumption. This combination offers a safe, effective, and practical enhancement to regional anesthesia for upper limb surgeries.

**Trial registration:** PACTR202509579633417 (retrospectively registered on 11 September 2025)

## Background

Pain is not merely a sensory modality but a multidimensional experience. The International Association for the Study of Pain (IASP) defines it as “an unpleasant sensory and emotional experience associated with actual or potential tissue damage” [1]. This definition highlights the interplay between subjective perception, emotional response, and physiological mechanisms. Pain response vary widely among individuals and even within the same person over times, influenced by factors such as age, gender and psychological state. In hospital settings, both acute and chronic pain are common, making postoperative pain management essential for all patients [1].

Despite advances in postoperative pain (POP) management, significant gaps remain. Acute POP is often undertreated, particularly following general surgical procedures. Unrelieved pain can negatively affect physical and psychological well-being, delay recovery, and increase healthcare costs. Immediate consequences of severe pain include reduced mobility, sleep disturbances, increased reliance on analgesics, and impaired immune function, which may heighten susceptibility to infection [2]. Poorly managed pain can also trigger pathophysiological changes in the peripheral and central nervous systems, potentially leading to chronic pain. In such cases, central sensitization may result in exaggerated pain responses to stimuli that would otherwise be tolerable. [3].

Regional anesthesia offers excellent analgesia and is widely accepted for POP relief [4]. For upper limb surgeries, brachial plexus block (BPB) is used either as an adjunct to general anesthesia or as a sole anesthetic technique. However, the duration of analgesia is limited by the pharmacokinetics of local anesthetics (LAs). Prolongation can be achieved through indwelling perineural catheters or the addition of adjuvant drugs. While catheter techniques provide extended analgesia, they are limited by technical challenges such as placement accuracy, risk of infection, and removal difficulties [5].

Recent advancements in peripheral nerve block (PNB) techniques and the use of adjuvants have significantly improved analgesic outcomes. Adjuvants—agents administered alongside LAs—enhance block quality, extend analgesia duration, and improve patient recovery [6]. A comprehensive understanding of the mechanisms of action, clinical effects, and safety profiles of PNB and the use of additional adjuvants is essential.

Brachial plexus blocks can be performed via interscalene, supraclavicular, infraclavicular, or axillary approaches. Among these, the supraclavicular route is preferred for procedures below the shoulder due to its consistency and ease of access. However, complications such as pneumothorax, hemothorax, Horner’s syndrome, and phrenic nerve block may occur. Ultrasound-guided supraclavicular blocks improve safety by allowing real-time visualization of anatomical structures and needle placement.

Although LAs alone provide good intraoperative conditions, their effects wear off within hours, exposing patients to moderate to severe postoperative pain [7]. To address this, various adjuvants—including morphine, tramadol, fentanyl, clonidine, dexmedetomidine, and midazolam—have been used. [8]. Recently, corticosteroids such as dexamethasone have gained attention for their analgesic and antiemetic properties. Dexamethasone can be administered systemically or perineurally and has shown efficacy in enhancing peripheral nerve blocks [9].

Dexamethasone is a long-acting glucocorticoid (half-life >36 hours) with potent anti-inflammatory and analgesic effects [10]. It prolongs nerve block duration by inhibiting nociceptive C-fiber transmission and modulating potassium channel activity in excitable cells [11]. Additionally, dexamethasone induces vasoconstriction, slowing local anesthetic absorption and extending its effect [12].

A single 8 mg dose of intravenous dexamethasone combined with bupivacaine in supraclavicular brachial plexus block has been shown to reduce postoperative pain intensity and prolong analgesia without adverse effects. Its low cost, antiemetic properties, and favorable safety profile make it a practical alternative to perineural administration, especially in resource-limited settings [13]. To date, no studies have investigated this approach in Eritrea, and research on this topic remains scarce. Therefore, this study was conducted to compare the duration of analgesia between two groups—one receiving bupivacaine alone and the other receiving bupivacaine with intravenous dexamethasone—in supraclavicular brachial plexus blocks.

## Materials & Method

### Study design & setting

This study was a hospital-based, randomized clinical trial designed to evaluate the effect of intravenous dexamethasone combined with bupivacaine on prolonging postoperative analgesia following supraclavicular brachial plexus block (SCBPB). It was conducted in the orthopedic department of Halibet National Referral Hospital (HNRH), the sole government-run tertiary center providing orthopedic services in Eritrea. The hospital is located in Asmara, the capital city.

### Study participants

Sixty adult patients scheduled for elective upper limb surgeries under SCBPB were enrolled. Inclusion criteria were: age between 18 and 80 years, ASA physical status I or II, and procedures involving the elbow, forearm, or hand. Exclusion criteria included: patient refusal, contraindications to SCBPB (e.g., severe chronic obstructive pulmonary disease, contralateral diaphragmatic paralysis, neuropathy of the surgical limb), pregnancy, bleeding disorders, local infection at the injection site, known allergy to local anesthetics, long-term steroid therapy, use of premedications that could affect outcomes (opioids, ketamine, NSAIDs), diabetes mellitus, hepatic or renal disease.

### Sample size

Sample size was calculated using the formula for comparing means of quantitative outcomes, accounting for Type I error (α = 0.05, Zα = 1.96) and Type II error (β = 0.10, power = 90%, Zβ = 1.28). Based on data from a similar study, the variances were:

σ₁² (dexamethasone group) = 180.04

σ₂² (control group) = 110.62

μ₁ = 1089

μ₂ = 364.52 [2].

The minimum required sample size was 7 per group. To enhance robustness, 30 participants were included in each group.

### Research variables

Primary Outcome: Duration of postoperative analgesia,

Secondary Outcome: Onset of sensory and motor block, Postoperative pain scores, Analgesic consumption

Independent variables: Perineural Bupivacaine, intravenous dexamethasone injection, intravenous normal saline

Patient variables: Socio-demographic & clinical characteristics.

### Data Collection and Anesthesia Protocol

Sociodemographic and clinical data (age, sex, education, religion, ethnicity, weight, height, ASA status, BMI, prior anesthesia/surgery history, analgesic doses and timing) were collected using a structured checklist. A standard pre-anesthetic assessment was performed, and informed consent obtained.

The data collection started on September 23, 2024 and ended on January 17, 2025.

All SCBPBs were performed by anesthetists with at least two years of experience in ultrasound-guided techniques. A designated anesthetist administered the block independently. After drug administration, vital signs were recorded, and sensory/motor block onset was assessed every 2 minutes for 15 minutes.

Sensory block: Assessed via pinprick on a 3-point scale (2 = normal sensation, 1 = loss of pinprick sensation, 0 = loss of light touch).

Motor block: Assessed using the modified Bromage scale (2 = complete block, 1 = reduced strength, 0 = normal function).

Vital signs were monitored every 10 minutes until the end of surgery.

Postoperative pain intensity, time to first analgesic request, type and total analgesic use within 24 hours were recorded. Patients were instructed to self-report pain using the Numeric Rating Scale (NRS), a 10 cm line where 0 = no pain and 10 = worst imaginable pain. Pain scores were recorded at 1, 2, 4, 6, 8, 12, and 24 hours postoperatively. Adverse events (nausea, vomiting, seizures, hypotension, bradycardia, respiratory depression, allergic reactions) were documented. Data completeness and consistency were verified.

### Randomization and Blinding

Participants were randomly assigned using a simple lottery method due to limited daily surgical volume. Allocation concealment was maintained with sealed opaque envelopes. Researchers generated the sequence, enrolled participants, and assigned interventions. Both participants and outcome assessors were blinded; the anesthetist performing the block was not involved in assessments.

### Intervention

Ethical approval was obtained from the Orotta College of Medicine and Health Sciences Ethics Committee and the Ministry of Health’s Department of Research and Human Resources. The study adhered to CONSORT guidelines. Additional permission was granted by the hospital administration.

Participants were randomly assigned to two groups using a simple lottery method: Experimental group: Received 20 ml of 0.5% bupivacaine plus 8 mg IV dexamethasone Control group: Received 20 ml of 0.5% bupivacaine alone

Postoperative monitoring and pain assessments were conducted in the ward at seven time points (1st, 2nd, 4th, 6th, 8th, 12th, and 24th hours). NRS scores and checklist data were used to evaluate pain. Patients requiring additional analgesia or experiencing complications were managed accordingly.

### Data Analysis

Data were entered into SPSS version 26.0 (IBM Corp., Chicago, USA). After cleaning, descriptive and inferential statistics were performed. Categorical variables were analyzed using frequencies, percentages, and chi-square tests. Quantitative variables were tested for normality using the Shapiro-Wilk test. Normally distributed variables were summarized using means and standard deviations and compared using independent samples t-tests. Non-normally distributed variables were summarized using medians and interquartile ranges and compared using Mann-Whitney U tests. A p-value < 0.05 was considered statistically significant.

### CONSORT Flowchart

## Results

### Sociodemographic characteristics of the patients

The sociodemographic characteristics of the patients are shown in Table 1. A total of 60 patients were enrolled. The median age was 26.50 (IQR = 23), ranging from 18 to 80 years. Female participants comprised 26.67% of both the control and experimental groups. Similarly, the distribution of the patients by educational level was not significantly different between the control and experimental groups, with most patients having completed secondary education (41.67%).

**Table 1:**
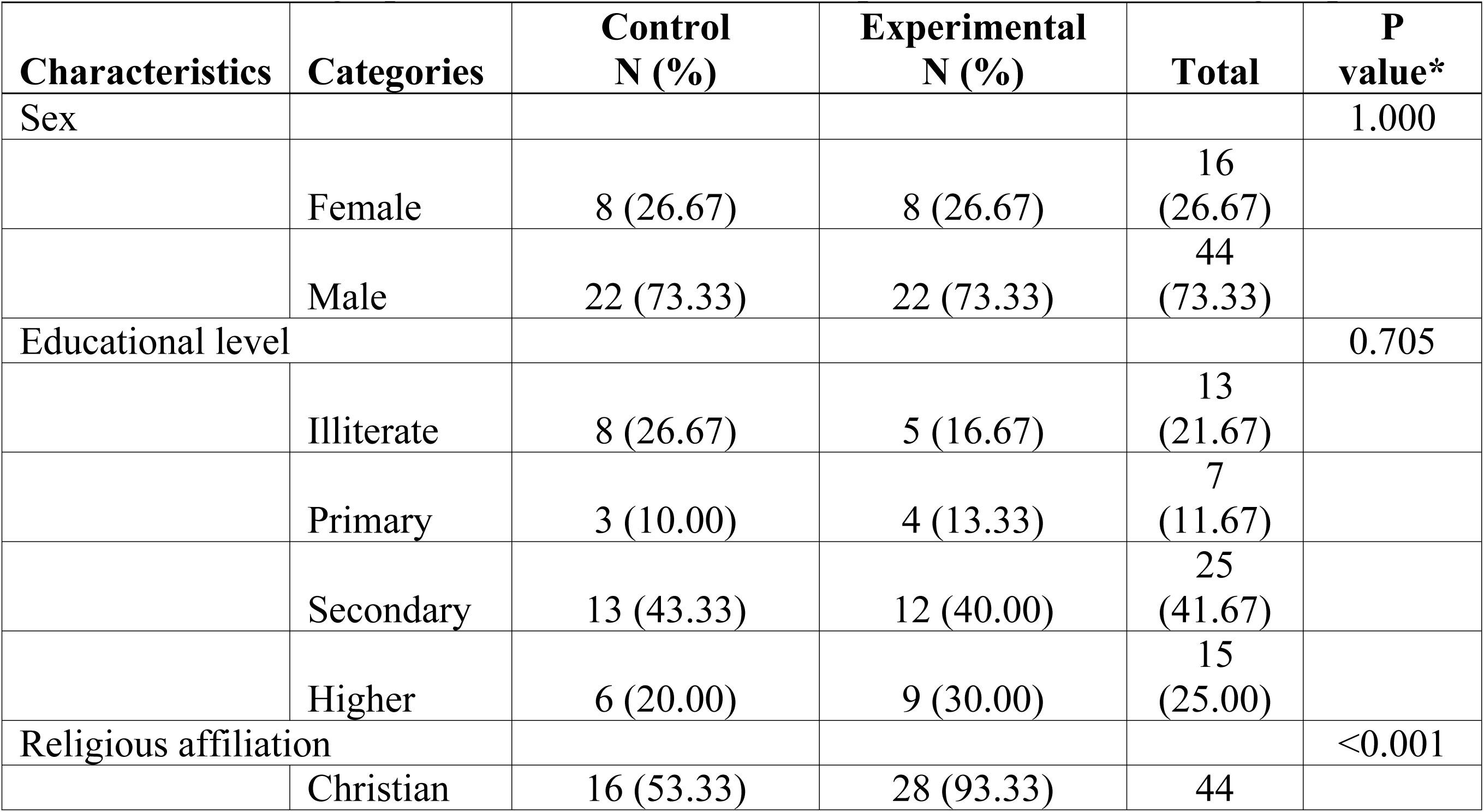
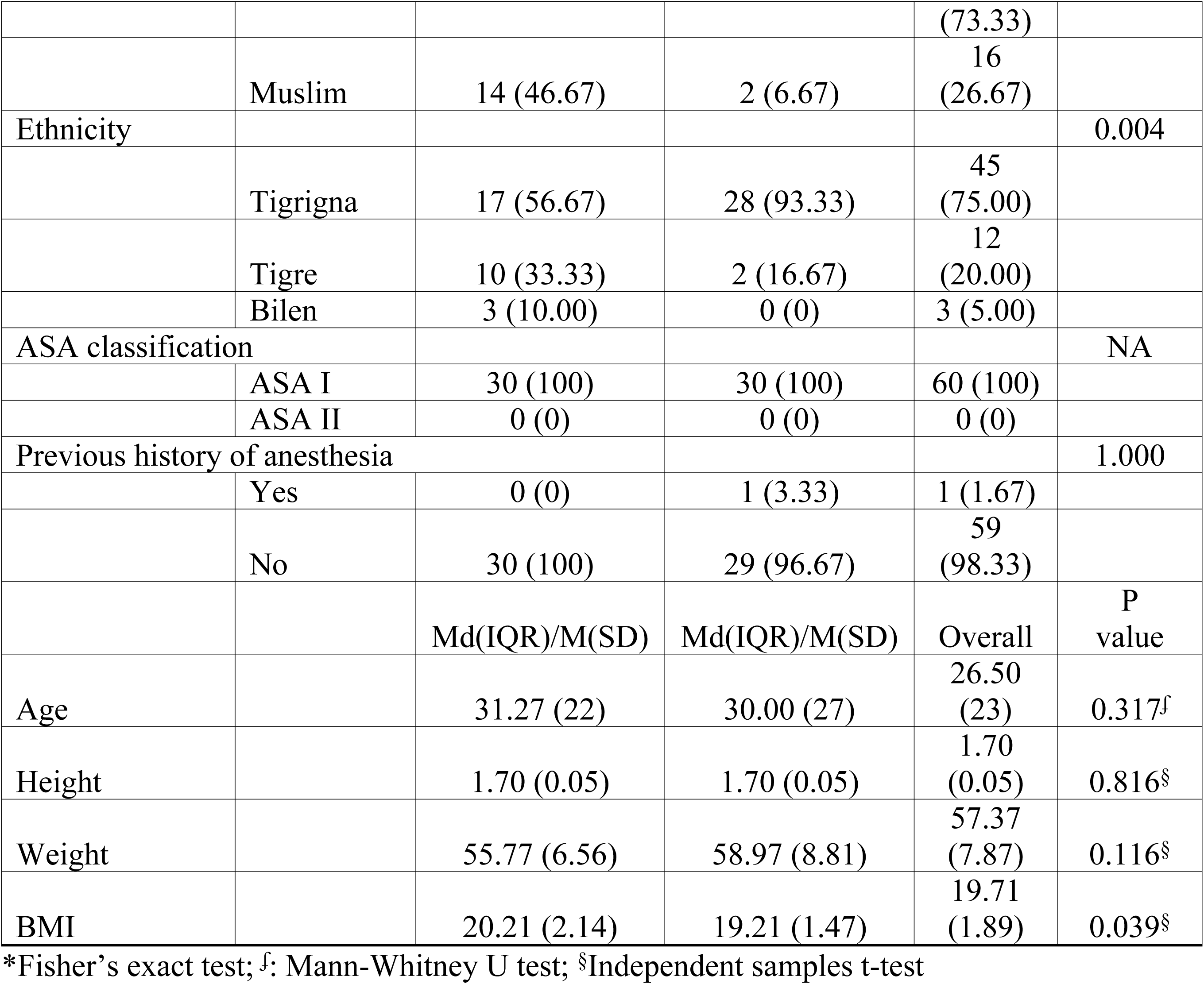
Sociodemographic characteristics of the experimental and control groups.

However, religious affiliation and ethnicity differed significantly between groups. The experimental group had a higher proportion of Christians (93.33%) compared to the control group (53.33%) (p < 0.001). Similarly, Tigrigna ethnicity was more prevalent in the experimental group (93.33%) than in the control group (56.67%) (p = 0.004). All patients were ASA I. Only one patient in the experimental group had a history of general anesthesia, and nine had received premedication.

No significant differences were found in age (p = 0.317), height (p = 0.816), or weight (p = 0.116) between groups. However, BMI was significantly lower in the experimental group (M = 19.21, SD = 1.47) compared to the control group (M = 20.21, SD = 2.14) (p = 0.039).

### Preoperative vital signs

The preoperative vital signs of the patients were assessed and are reported in Table 2. Baseline vital signs were comparable between groups. No significant differences were observed in heart rate (p = 0.204), systolic blood pressure (p = 0.782), diastolic blood pressure (p = 0.228), respiratory rate (p = 0.817), or oxygen saturation (SpO₂) (p = 0.702).

**Table 2:**
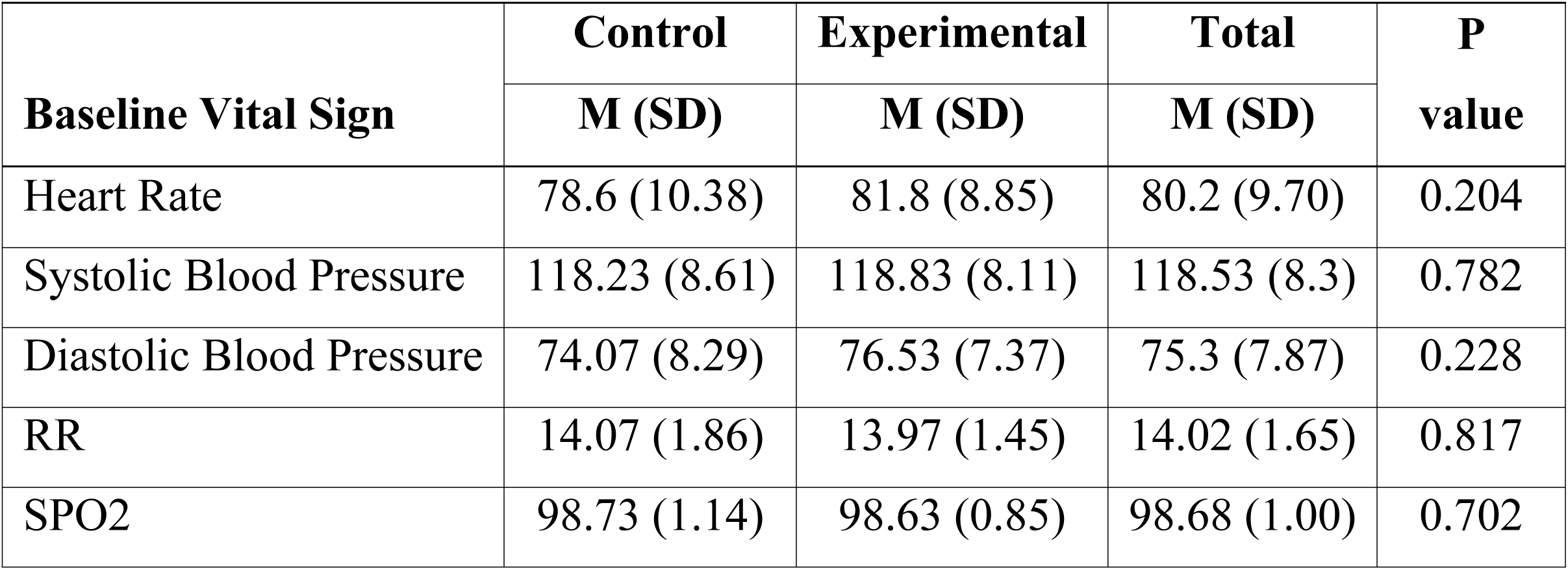
Comparison of the pre-operative vital sign of the patients in the two groups.

**Table 3:** Comparison between the control and experimental groups according to HR, SBP, DBP, RR, and SpO2.

| Vital Sign |  | Control<br>Mean ± SD | Experiment<br>Mean ± SD | MD (95% CI) | P<br>Value |
| --- | --- | --- | --- | --- | --- |
| Heart Rate |  |  |  |  |  |
|  | 5 minutes | 77.6 ± 8.4 | 79.1 ± 9.43 | -1.5 (-6.11, 3.11) | 0.518 |
|  | 15 minutes | 77.43 ± 7.94 | 76.67 ± 6.19 | 0.77 (-2.91, 4.45) | 0.678 |
|  | 25 minutes | 76.1 ± 5.96 | 76.4 ± 6.74 | -0.3 (-3.59, 2.99) | 0.856 |
|  | 35 minutes | 75.57 ± 5.73 | 74.6 ± 3.57 | 0.97 (-1.5, 3.43) | 0.436 |
|  | 45 minutes | 75.27 ± 6.15 | 74.23 ± 3.8 | 1.03 (-1.61, 3.68) | 0.437 |
|  | 55 minutes | 72.73 ± 3.53 | 74.07 ± 3.76 | -1.33 (-3.22, 0.55) | 0.162 |
| Systolic Blood Pressure |  |  |  |  |  |
|  | 5 minutes | 118.57 ± 6.98 | 118.37 ± 6.6 | 0.2 (-3.31, 3.71) | 0.910 |
|  | 15 minutes | 118.8 ± 5.83 | 119.47 ± 5.36 | -0.67 (-3.56, 2.23) | 0.647 |
|  | 25 minutes | 118.73 ± 6.62 | 114.7 ± 20.12 | 4.03 (-3.71, 11.77) | 0.301 |
|  | 35 minutes | 119.67 ± 4.15 | 119.2 ± 4.56 | 0.47 (-1.79, 2.72) | 0.680 |
|  | 45 minutes | 118.37 ± 4.7 | 117.33 ± 3.08 | 1.03 (-1.02, 3.09) | 0.318 |
|  | 55 minutes | 118.7 ± 3.67 | 118.1 ± 3.8 | 0.6 (-1.33, 2.53) | 0.536 |
| Diastolic Blood Pressure |  |  |  |  |  |
|  | 5 minutes | 75.77 ± 5.12 | 74.47 ± 5.73 | 1.3 (-1.51, 4.11) | 0.358 |
|  | 15 minutes | 77.2 ± 5.09 | 77.8 ± 4.14 | -0.6 (-3, 1.8) | 0.618 |
|  | 25 minutes | 77.87 ± 5.34 | 77.4 ± 5.12 | 0.47 (-2.23, 3.17) | 0.731 |
|  | 35 minutes | 78.97 ± 4.88 | 77.47 ± 4.17 | 1.5 (-0.85, 3.85) | 0.206 |
|  | 45 minutes | 77.67 ± 3.6 | 77.37 ± 4.8 | 0.3 (-1.89, 2.49) | 0.785 |
|  | 55 minutes | 75.33 ± 13.3 | 77.27 ± 4.83 | -1.93 (-7.1, 3.24) | 0.457 |
| RR |  |  |  |  |  |
|  | 5 minutes | 14 ± 1.8 | 14 ± 2.84 | 0 (-1.23, 1.23) | 1.00 |
|  | 15 minutes | 14.1 ± 1.6 | 14 ± 2.3 | 0.1 (-0.93, 1.13) | 0.846 |
|  | 25 minutes | 13.73 ± 1.23 | 13.83 ± 1.49 | -0.1 (-0.81, 0.61) | 0.778 |
|  | 35 minutes | 13.9 ± 1.69 | 13.73 ± 1.57 | 0.17 (-0.68, 1.01) | 0.694 |
|  | 45 minutes | 13.73 ± 1.34 | 13.63 ± 1.83 | 0.1 (-0.73, 0.93) | 0.810 |
|  | 55 minutes | 14.17 ± 1.93 | 13.73 ± 2.03 | 0.43 (-0.59, 1.46) | 0.401 |
| SPO<sub>2</sub> |  |  |  |  |  |
|  | 5 minutes | 99 ± 1.14 | 98.8 ± 1.03 | 0.2 (-0.36, 0.76) | 0.480 |
|  | 15 minutes | 99.1 ± 0.8 | 98.57 ± 0.94 | 0.53 (0.08, 0.98) | **0.021** |
|  | 25 minutes | 99.27 ± 0.94 | 99.03 ± 0.81 | 0.23 (-0.22, 0.69) | 0.308 |
|  | 35 minutes | 99.23 ± 0.86 | 99.33 ± 0.71 | -0.1 (-0.51, 0.31) | 0.625 |
|  | 45 minutes | 99.03 ± 1.22 | 98.97 ± 1.25 | 0.07 (-0.57, 0.7) | 0.835 |
|  | 55 minutes | 98.8 ± 1.13 | 98.57 ± 1.04 | 0.23 (-0.33, 0.79) | 0.408 |

**Table 4:** Comparison of the duration of supraclavicular brachial plexus block.

|  | Control | Experimental |  |
| --- | --- | --- | --- |
| Variable | Mean ± SD | Mean ± SD | P value |
| Duration of surgery, in minutes | 64.60 ± 14.10 | 67.33 ± 16.75 | 0.497 |
| Duration of analgesia, in hours | 7.57 ± 1.04 | 17.36 ± 3.75 | <0.001 |
| Onset of sensory block, in minutes | 12.03 ± 2.39 | 6.53 ± 1.04 | <0.001 |
| Onset of motor block, in minutes | 15.37 ± 1.96 | 9.63 ± 1.38 | <0.001 |

**Table 5.** : **Postoperative pain severity scores between the groups**

| Time | Control | Level | Experimental | Level | P |
| --- | --- | --- | --- | --- | --- |

|  |  |  |  |  | value |
| --- | --- | --- | --- | --- | --- |
| At 1st hour | 0 (0) | No pain | 0 (0) | No pain | 1.000 |
| At 2nd hour | 0 (0) | No pain | 0 (0) | No pain | 1.000 |
| At 4th hour | 1.77 (0.43) | Mild pain | 0.23 (0.43) | No pain | <0.001 |
| At 6th hour | 3.17 (0.91) | Mild pain | 1.2 (0.48) | Mild pain | <0.001 |
| At 8th hour | 4.68 (0.75) | Moderate pain | 2.2 (0.48) | Mild pain | <0.001 |
| At 12th hour | 5 (0) | Moderate pain | 3.43 (0.9) | Mild pain | <0.001 |
| At 24th hour | - | - | 4.82 (0.58) | Moderate pain | - |

**Table 6:**
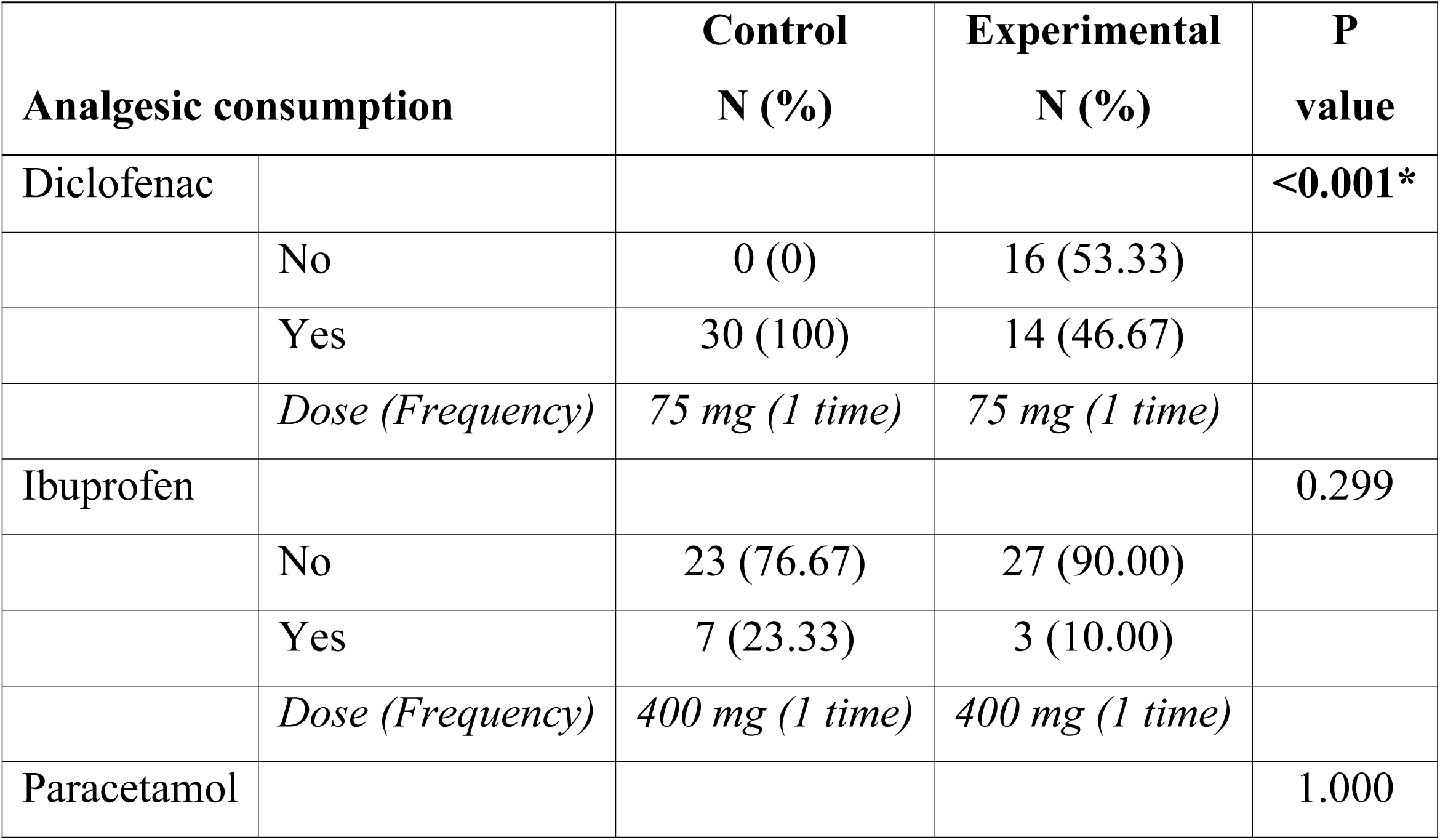
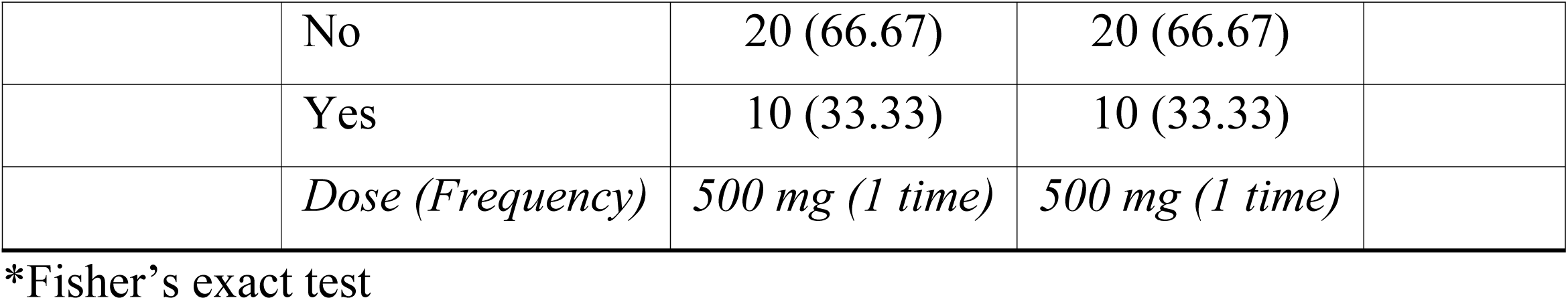
Requirement of analgesics in the two groups.

### Intraoperative Vital Signs

Intraoperative monitoring showed no significant differences in heart rate, systolic or diastolic blood pressure, or respiratory rate at any recorded time point. However, SpO₂ was significantly higher in the control group at 15 minutes (M = 99.1, SD = 0.8) compared to the experimental group (M = 98.57, SD = 0.94) (p = 0.021).

### Block Characteristics and Analgesia Duration

The duration of surgery was similar between groups (control: M = 64.60 min, SD = 14.10; experimental: M = 67.33 min, SD = 16.75; p = 0.497). However, the duration of analgesia was significantly longer in the experimental group (M = 17.36 hours, SD = 3.75) than in the control group (M = 7.57 hours, SD = 1.04) (p < 0.001). The results are graphically depicted in Figure 1.

**Figure 1.**
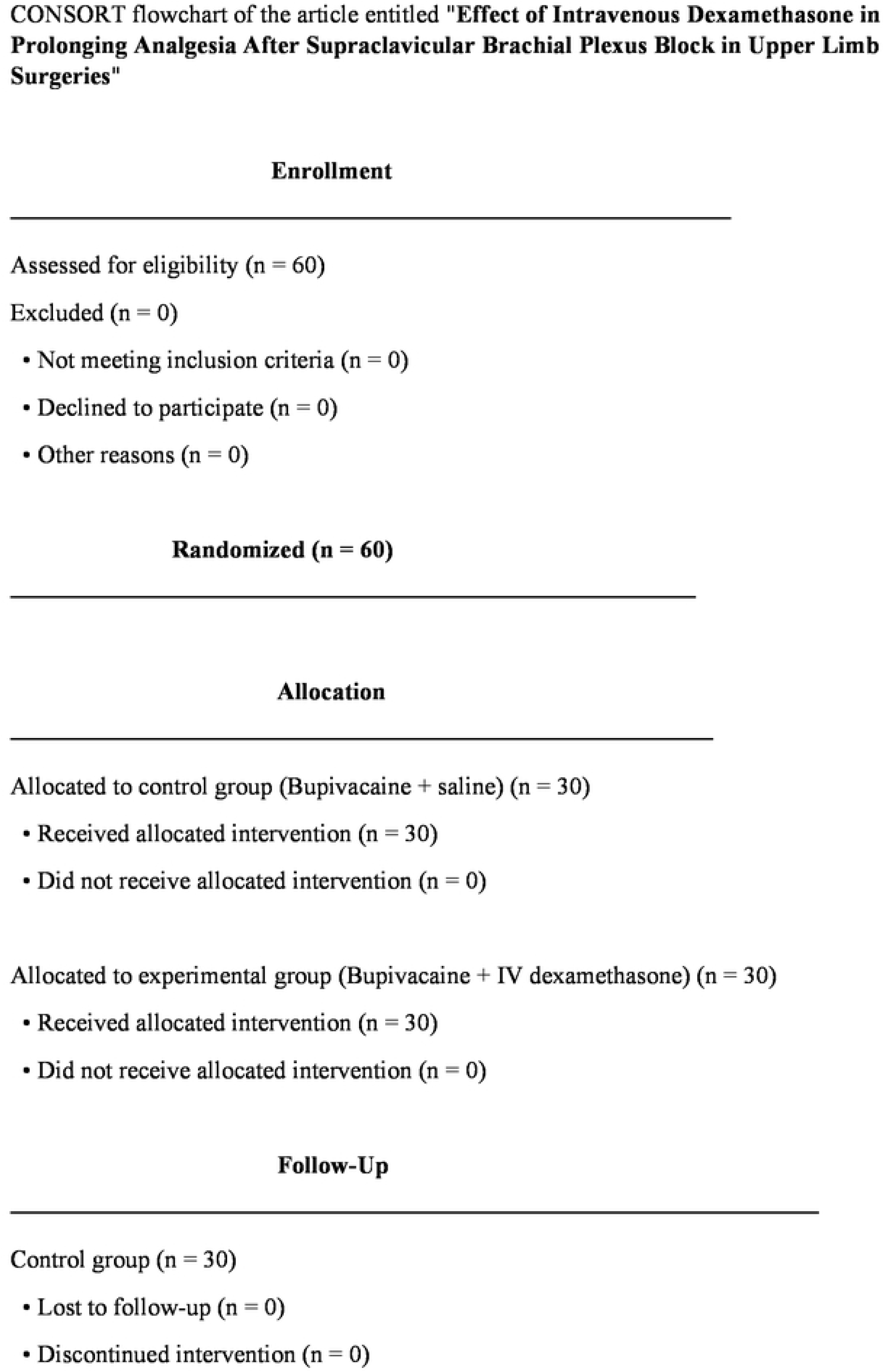
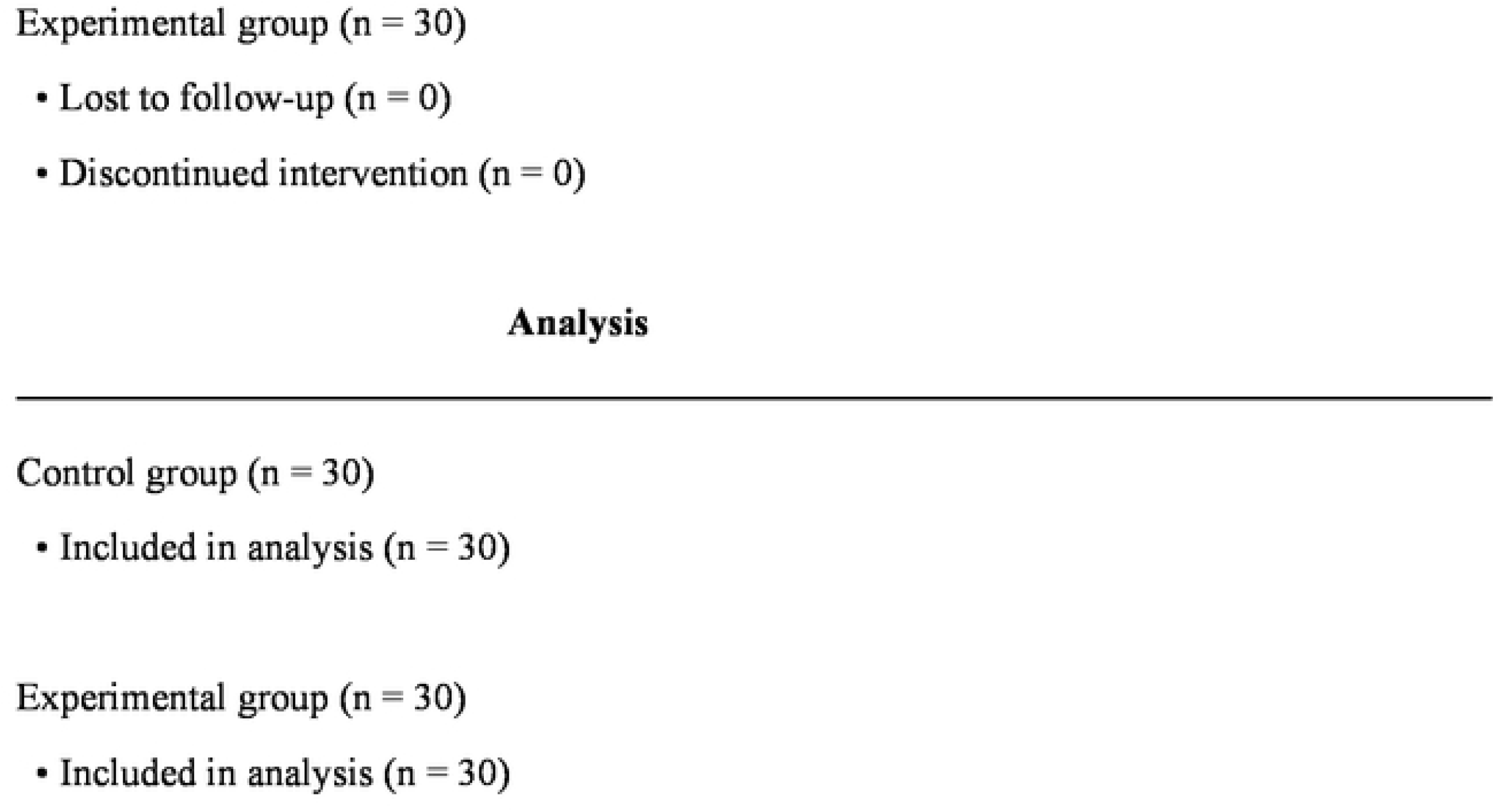
CONSORT Flowchart of the participants

**Figure 2:**
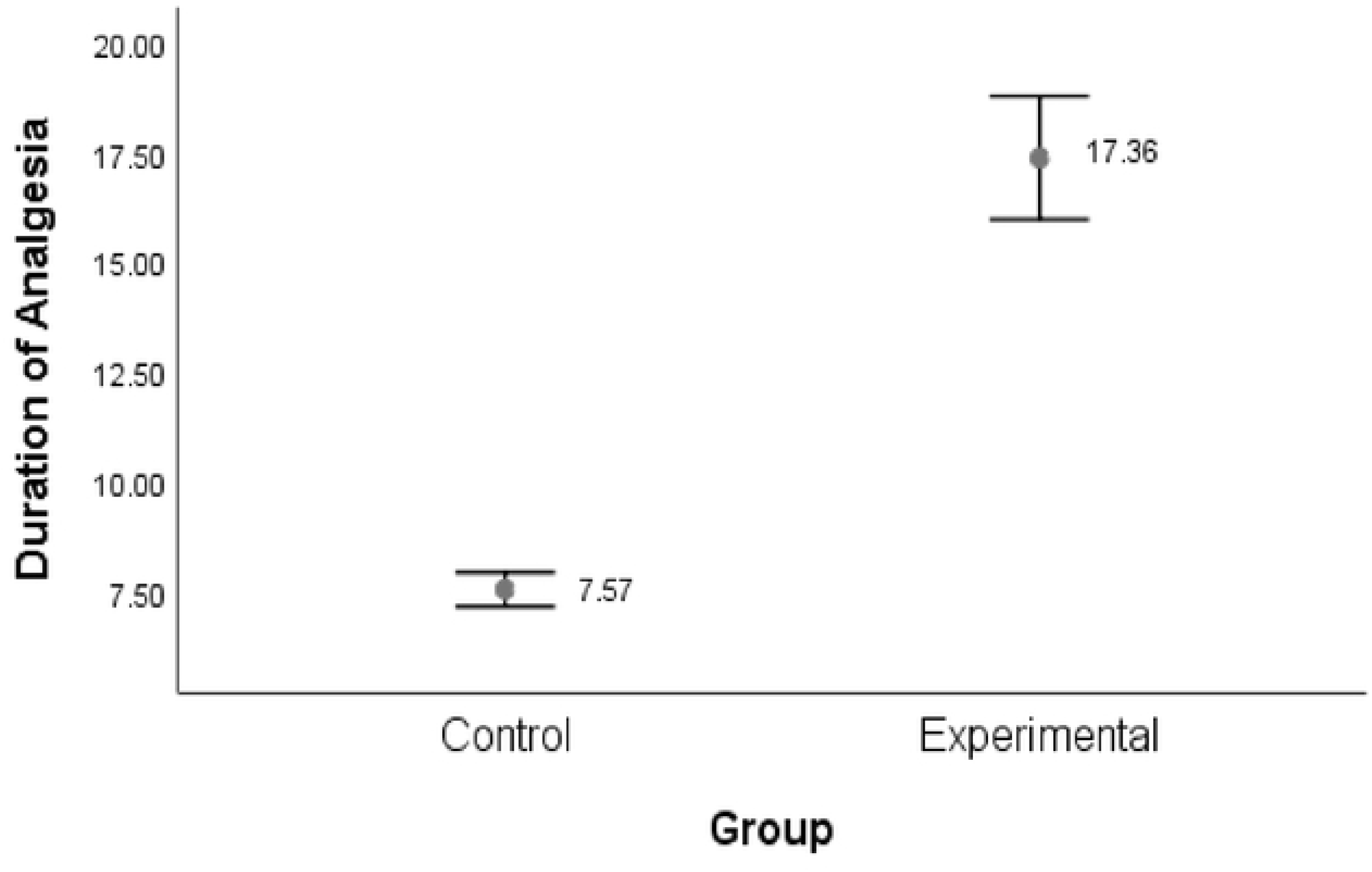
Comparison of the duration of analgesia in hours between the control and experimental groups

The onset of sensory block was faster in the experimental group (M = 6.53 min, SD = 1.04) compared to the control group (M = 12.03 min, SD = 2.39) (p < 0.001). Similarly, onset of motor block was faster in the experimental group (M = 9.63 min, SD = 1.38) than in the control group (M = 15.37 min, SD = 1.96) (p < 0.001).

### Postoperative Pain Scores

Postoperative pain was assessed using the Numeric Rating Scale (NRS) at seven time points. No significant differences were observed at the 1st and 2nd hours (p = 1.000). However, pain scores were significantly lower in the experimental group at the 4th (p < 0.001), 6th (p < 0.001), 8th (p < 0.001), and 12th hours (p < 0.001). At the 24th hour, pain scores remained lower in the experimental group, though not statistically tested.

A graphical depiction of the comparison of the postoperative mean NRS scores is shown in Figure 3. The experimental line was below the control line, indicating a lower postoperative mean NRS score. In addition, the error bars have no intersection revealing significant differences at 4^th^ hour, 6^th^ hour, 8^th^ hour, and 12^th^ hour.

**Figure 3:**
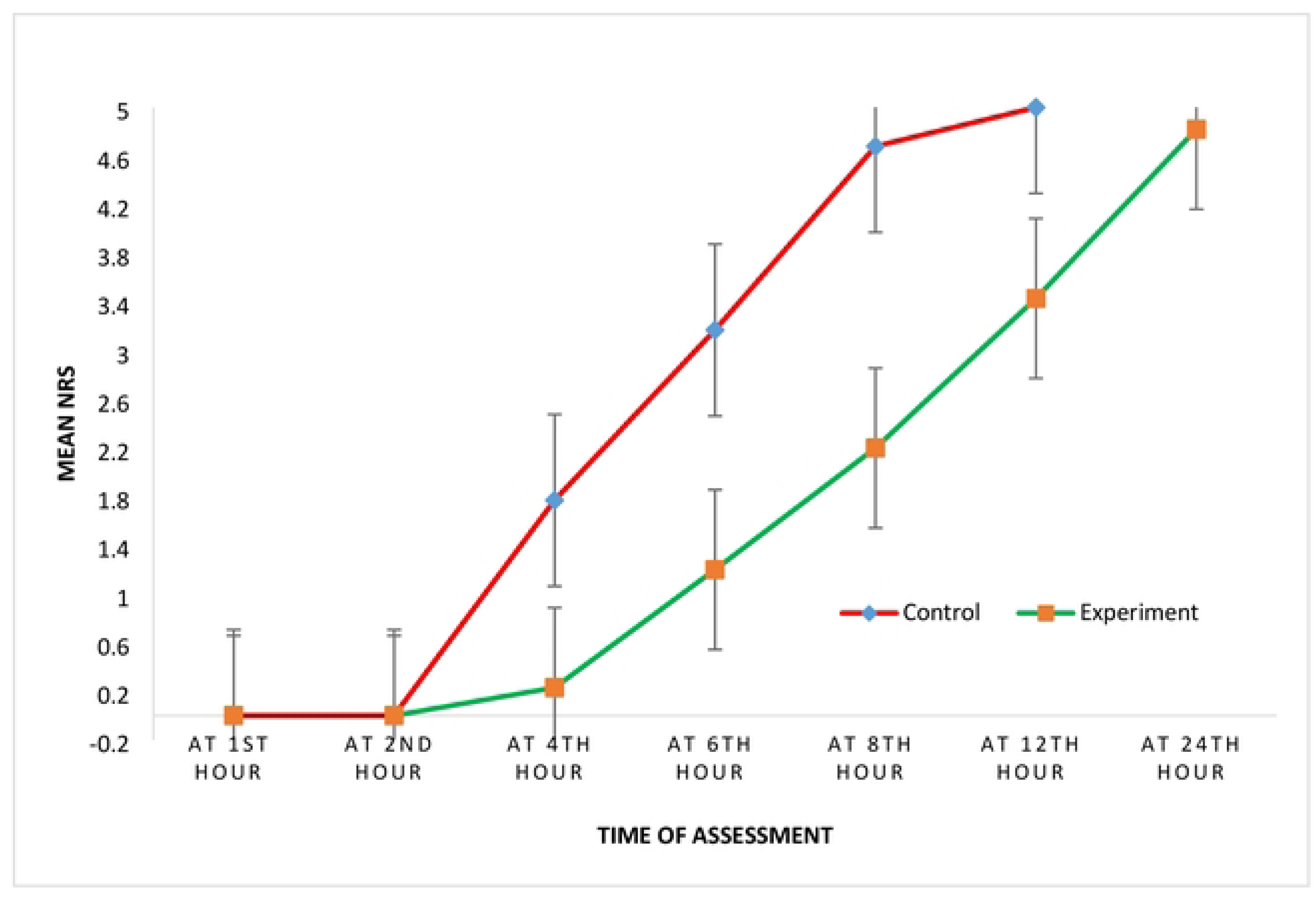
Comparison of postoperative pain severity scores between the groups

### Analgesic Consumption

Diclofenac use was significantly higher in the control group (100%) compared to the experimental group (46.67%) (p < 0.001). Paracetamol use was equal in both groups (33.33%). Ibuprofen use was not significantly different (control: 23.33%, experimental: 10.00%; p = 0.299). All analgesics were administered in standard doses and frequencies.

## Discussion

Supraclavicular brachial plexus block (SCBPB) has become a preferred technique among anesthetists for upper limb surgeries due to its ability to avoid complications associated with general anesthesia, particularly those related to airway instrumentation. It offers a safe, cost-effective, and efficient approach with excellent postoperative analgesia. This study aimed to evaluate the effects of intravenous dexamethasone combined with bupivacaine on the duration of analgesia, onset of sensory and motor block, and analgesic consumption in SCBPB.

Previous studies have shown that preoperative administration of dexamethasone—either orally or intravenously—can reduce postoperative pain scores and analgesic requirements [14]. In our trial, both groups were comparable in terms of age, sex, weight, ASA classification, surgical site, and duration of surgery (p > 0.05), consistent with findings from similar studies [15]. Hemodynamic parameters remained stable across both groups [16].

Our results demonstrated that intravenous administration of 8 mg dexamethasone significantly prolonged postoperative analgesia. Patients in the dexamethasone group experienced a mean analgesia duration of 17.36 hours (SD = 3.75), compared to 7.57 hours (SD = 1.04) in the control group—an approximate twofold increase. These findings align with prior studies reporting extended analgesia durations of 18–25 hours with dexamethasone. A study from Bangladesh also reported prolonged pain-free intervals, though shorter than ours, possibly due to differences in pain perception, cultural factors, and treatment protocols [17].

Another study performed in Egypt also revealed that the mean duration of analgesia in the dexamethasone group was 18.45±2.26 hours, whereas in the bupivacaine alone group, it was 10.33±1.54 hours (*p*=.001) [18]. Furthermore, our study was supported by a study performed in 2019, which revealed that the mean pain-free time was longer in the dexamethasone bupivacaine group than in the bupivacaine alone group with a p-value<0.0001 [19]. In contrast, a study performed in Bangladesh, reported that the first analgesic requirement time in the dexamethasone bupivacaine group was 864.50±25.19 minutes, whereas it was 455±17.09 minutes in the bupivacaine alone group [20]. The variance in pain perception evaluation and treatment, as well as differences in pain tolerance levels among societies, could be possible reasons for the shorter pain-free time. Genetics, social, and cultural factors, which vary among individuals, influence the pain experience.

Postoperative pain scores were significantly lower in the dexamethasone group at the 4th, 6th, 8th, and 12th hours. This may be attributed to the synergistic effect of dexamethasone with bupivacaine, which extends beyond the typical duration of bupivacaine alone. The faster onset of sensory and motor block in the dexamethasone group (6.53 min and 9.63 min, respectively) compared to the control group (12.03 min and 15.37 min) was also statistically significant.

In 2024, a study conducted in Nepal reported that the mean time onset of sensory blocks was 8.4 min in the BS group and 4.6 min in the BD group and the mean time onset of motor blocks was 13.2 min in the BS group and 8.4 min in the BD group [21]. In another study performed in Bangladesh, the onset of sensory block also lower in the dexamethasone group than in the plain local anesthetic group [22]. Therefore, the addition of dexamethasone to bupivacaine helps for the fast onset both on sensory and motor block. Dexamethasone enhances the potency of analgesic agents. This is also true when it comes to bupivacaine and it also provides a better postoperative prolonged analgesia.

Analgesic consumption within the first 24 hours was significantly lower in the dexamethasone group. Diclofenac use was notably higher in the control group, while paracetamol and ibuprofen usage showed no significant differences. These findings support previous research indicating that dexamethasone reduces the need for rescue analgesics and opioid consumption when administered at intermediate doses [23].

The mean number of rescue analgesic doses was significantly lower in the dexamethasone group than in the bupivacaine group. Similarly, in a study performed in Nepal, the mean number of rescue analgesic doses was also lower in the dexamethasone group than in the other groups [24].

Many previous studies have demonstrated that a single perioperative dose of dexamethasone is not associated with a significant increase in the incidence of adverse effects in either pediatric or adult patients [13]. In the present study, no patients experienced nausea or vomiting and there was no delay in readiness to discharge.

### Limitations of the study

Pain is inherently subjective and influenced by individual genetic, social, and cultural factors, which may affect the generalizability of our findings. Additionally, variability in pain tolerance and reporting may introduce bias in pain score assessments. These factors should be considered when interpreting the results.

## Conclusion

The addition of 8 mg intravenous dexamethasone to 0.5% bupivacaine in supraclavicular brachial plexus blocks significantly prolonged postoperative analgesia, accelerated the onset of sensory and motor block, and reduced analgesic consumption. This combination offers a safe, effective, and practical enhancement to regional anesthesia for upper limb surgeries.

## Declarations

### List of abbreviations

ASA: American Society of Anesthesiologists
BS: Bupivacaine and normal saline
BD: Bupivacaine and dexamethasone
CI: Confidence Interval
ENT: Ear nose throat
DOA: Duration of action
GI: Gastrointestinal
HNRH: Halibet National Referral Hospital
IV: Intravenous
ISB: Interscalene Block
LAs: Local Anesthetics
NRS: Numerical Rating Scale
PADSS: Post-Anesthetic Discharge Scoring System
PNB: Peripheral Nerve Block
POP: Postoperative Pain
SCBPB: Supraclavicular Brachial Plexus Block
SPSS: Statistical Package for Social Sciences
VAS: Visual Analog Scale
RCT: Randomized Clinical Trial
USG: Ultrasonography

### Ethics approval and consent to participate

Ethical clearance and approval were obtained from the Ethical and Scientific Committee of the Orotta College of Medicine and Health Sciences as well as from the Health Research Proposal Review and Ethical Committee of the Ministry of Health. Permission was also granted from Halibet National Referral Hospital. Additionally, the study is registered retrospectively in the Pan African Clinical Trial Registry. This study also fulfills the requirement of any subject protection and does not harm principle of a clinical trial which adheres to the declaration of Helsinki. After the purpose of the study was explained and confidentiality and anonymity was assured, informed written consent was obtained from each participant. Names and other identifying information were not included in the study. All methods were performed in accordance with relevant guidelines and regulations.

### Consent for publication

Not applicable

### Availability of data and materials

The datasets generated and/or analyzed during the current study are available from the corresponding author on reasonable request.

### Generalizability

This study supports the use of dexamethasone as an adjuvant in low-resource settings, offering prolonged analgesia and reduced burden on healthcare systems.

### Data Sharing

De-identified data will be shared upon reasonable request or when applicable.

### Trial Status

Recruitment and data collection are complete.

## Authors’ contributions

**MBM**: Study conception and design of the study, analysis and interpretation of the data, drafting, critical revision of the manuscript for important intellectual content, and submission; **SAH, AME, SHI** and **KHA**: Collection, acquisition, analysis and interpretation of the data; critical revision of the manuscript for important intellectual content

## Acknowledgement

The authors would like to thank the study participants for their participation in this study.

